# The Prevalence of Mental Health Disorders in people with HIV and the effects on the HIV Care Continuum

**DOI:** 10.1101/2022.04.19.22273931

**Authors:** Raynell Lang, Brenna Hogan, Jiafeng Zhu, Kristen McArthur, Jennifer Lee, Peter Zandi, Paul Nestadt, Michael J. Silverberg, Angela M. Parcesepe, Judith A. Cook, M. John Gill, David Grelotti, Kalysha Closson, Viviane D. Lima, Joseph Goulet, Michael A. Horberg, Kelly A. Gebo, Reena M. Camoens, Peter F. Rebeiro, Ank E. Nijhawan, Kathleen McGinnis, Joseph Eron, Keri N. Althoff, the North American AIDS Cohort Collaboration on Research and Design (NA-ACCORD) of the International Epidemiologic Databases to Evaluate AIDS (IeDEA)

**Author notes:** **CORRESPONDING AUTHOR:** Keri N Althoff, MPH, PhD, Associate Professor, Department of Epidemiology, Johns Hopkins Bloomberg School of Public Health, 615 N Wolfe Street, #E7142, Baltimore, MD, 21205.

## Abstract

**Objective:** To describe the prevalence of diagnosed depression, anxiety, bipolar disorder, and schizophrenia in people with HIV (PWH) and the differences in HIV care continuum outcomes in those with and without mental health disorders (MHD).

**Design:** Observational study of participants in the NA-ACCORD.

**Methods:** PWH (≥18 years) contributed data on prevalent schizophrenia, anxiety, depressive, and bipolar disorders from 2008-2018 based on ICD code mapping. MH multimorbidity was defined as having ≥ 2 MHD. Log binomial models with generalized estimating equations estimated adjusted prevalence ratios (aPR) and 95% confidence intervals for retention in care (≥ 1 visit/year) and viral suppression (HIV RNA ≤ 200 copies/mL) by presence vs. absence of each MHD between 2016-2018.

**Results:** Among 122,896 PWH, 67,643 (55.1%) were diagnosed with ≥ 1 MHD: 39% with depressive disorders, 28% with anxiety disorders, 10% with bipolar disorder, and 5% with schizophrenia. The prevalence of depressive and anxiety disorders increased between 2008-2018, while bipolar disorder and schizophrenia remained stable. MH multimorbidity affected 24% of PWH. From 2016-2018 (N=64,684), retention in care was marginally lower among PWH with depression or anxiety, however those with MH multimorbidity were more likely to be retained in care. PWH with bipolar disorder had marginally lower prevalence of viral suppression (aPR=0.98 [0.98-0.99]) as did PWH with MH multimorbidity (aPR=0.99 [0.99-1.00]) compared with PWH without MHD.

**Conclusion:** The prevalence of MHD among PWH was high, including MH multimorbidity. Although retention and viral suppression were similar to people without MHD, viral suppression was lower in those with bipolar disorder and MH multimorbidity.

## INTRODUCTION

With advances in HIV care, chronic disease management including care for co-occurring mental health disorders (MHD) among people with HIV (PWH) has become a primary focus of attention^[1]^. MHD remain a significant source of morbidity and mortality across the world;^[2]^ however, health outcomes of PWH with MHD remains under-studied, particularly within the Treat-All era (2015 and later)^[3]^. At the introduction of highly active antiretroviral therapy (ART) (in 1996), the first national estimates of 12-month prevalence of MHD among PWH in the US reported that nearly half of PWH screened positive for ≥ 1 of the following: major depression, dysthymia, generalized anxiety disorder, or panic attacks^[4]^. More recent studies have reported the prevalence of major depression, anxiety, bipolar disorder (BD), and schizophrenia are all more common among PWH compared to the general population^[5–13]^.

MHD have been associated with adverse outcomes in PWH, including poor adherence to ART, reduced viral suppression and excess mortality^[5–8,14]^. A correlation between depressive symptoms, lower CD4 counts and higher HIV RNA levels has been identified^[15]^. Prior to the Treat-All era, Moore et al. documented adherence to ART in PWH with BD to be 48% compared to 91% among PWH without BD^[16, 17]^. Schizophrenia in PWH has been associated with reduced linkage into care and poor adherence to treatment^[18]^. PWH who have effective treatment of their psychiatric symptoms are more successful in their HIV treatment^[9, 19]^. This association underlines the importance of early screening and evidence-based treatment for MHD among PWH[20].

HIV and depressive disorders (individually) are predicted to be the top two leading causes of burden of disease by 2030, with the greatest predicted years lost to disability^[21]^. Estimates of the burden of MHD among PWH are important for guiding policy and programs to ensure access to care, retention in care, viral suppression, and increased well-being of PWH. The objective of this study was to describe the prevalence of schizophrenia, anxiety, bipolar and depressive disorders in PWH in North America, and the effect of these disorders on the HIV care cascade during the transition to the Treat-All era.

## METHODS

### Study population

The North American AIDS Cohort Collaboration on Research and Design (NA-ACCORD) is a consortium of HIV cohort studies located throughout the United States and Canada, which was formed as part of the International epidemiology Databases to Evaluate AIDS (IeDEA) initiative of the National Institute of Allergy and Infectious Diseases^[22]^. Participants in contributing clinical cohorts had to link into care with ≥2 HIV care visits in 12 months to be eligible for enrollment in the NA-ACCORD. NA-ACCORD participants are demographically representative of the population of PWH in the US (according to US Centers of Disease Control and Prevention HIV surveillance data)^[23]^.

Each participating cohort annually submits data in a standardized format to the Data Management Core (DMC, University of Washington, Seattle WA). The DMC assesses data quality, harmonizes the data, and transfers it to the Epidemiology/Biostatistics Core (EBC, Johns Hopkins University, Baltimore MD). The EBC assesses the completeness of data elements and calculates cohort-specific “observation-windows” for specific outcomes to minimize the risk of falsely assuming complete event ascertainment from electronic health records^[24]^. Each participating cohort has been granted ethics approval by their respective local institutional review boards, as well as by the Johns Hopkins School of Medicine.

The source population for our nested study were PWH (≥18 years) participating in one of 13 NA-ACCORD-contributing clinical cohorts ascertaining anxiety, depression, BD, and schizophrenia diagnoses. Individual-level selection criteria for our study population included those who were observed in care from Jan 1, 2008 – Dec 31, 2018 (study period).

### Mental Health Disorders (MHD)

Cohorts collected inpatient and outpatient mental health diagnoses (and date at diagnosis) from health records using a list of ICD-9 and ICD-10 codes for depression, anxiety, BD, and schizophrenia (**Supplement Table 1**). The NA-ACCORD DMC mapped the ICD codes to create standardized definitions and the NA-ACCORD Mental Health Working group, in partnership with the NA-ACCORD EBC, established operationalized definitions of these MHD. Each MHD was defined as having at least one documented diagnosis over the follow-up period. A diagnosis of BD excluded classification of this participant as having depression; these two diagnoses were considered mutually exclusive^[25]^. Treatment data for BD and schizophrenia was obtained at the first time an individual met the criteria for a BD or schizophrenia diagnosis and was prescribed a medication indicated for treatment of these conditions (**Supplemental Table 1** outlines medications used to indicate treatment status). Due to the lack of specificity of medications for depression and anxiety, treatment data for these disorders were not included.

### Mental Health (MH) Multimorbidity

Mental health (MH) multimorbidity was defined as ≥2 mental health diagnoses (depression, anxiety, BD, or schizophrenia).

### HIV Care Continuum Outcomes: Retention in care and viral suppression

Retention in care was defined as having at least one in-person HIV primary care visit within a calendar year. Viral suppression was defined as having an HIV RNA <200copies/mL at the patient’s last measurement of the year.

### Covariates of interest

Sex was defined as sex at birth. Race/ethnicity was grouped into non-Hispanic white, non-Hispanic Black, Hispanic, and other/unknown. HIV acquisition risk group was determined at enrollment into the NA-ACCORD; using a mutually exclusive hierarchy: 1) injection drug use (IDU); 2) men who have sex with men (MSM); 3) heterosexual sexual contact; 4) other/unknown risk. Cigarette smoking was defined as ever reported use, via self-report or substance survey. At-risk alcohol use was defined as ever having an alcohol abuse or dependence diagnosis in the medical record. Hepatitis B (HBV) and Hepatitis C (HCV) infection were parameterized as ever/never. HBV was determined by a positive HBV surface antigen test, a positive HBV e-antigen test, or detectable HBV DNA. HCV was determined by a positive HCV antibody test, detectable HCV RNA, or the presence of any HCV genotype. CD4 count and HIV RNA were measured at baseline (i.e., the closest measurement to study entry, within 9 months prior through 3 months after study entry). ART regimen was also assessed at baseline (i.e., the closest record to study entry, ±6 months).

Comorbidities were assessed at baseline (i.e., prior to study entry, to 9 months after entry) using NA-ACCORD operationalized definitions. Treated hypertension was defined as having a hypertension diagnosis and prescription for antihypertensive medication^[26]^. Diabetes mellitus was defined as either: a type 2 diabetes diagnosis with prescription for diabetes-related medication; any prescription for diabetes-specific medication; or HgA1c ≥6.5%^[26]^. Chronic kidney disease (CKD) was defined as having eGFR calculated using the CKD-EPI study equation <60mL/min (Stage 2), based on serum creatinine measured closest to study entry^[26]^. Hypercholesterolemia was defined as having any total cholesterol measurements ≥240 mg/dL prior to statin initiation^[27]^.

### Statistical Analysis

#### Study entry

was defined as the date the cohort began observing patients, MH observation-window open date, patient enrollment into the NA-ACCORD, or January 1, 2008, whichever came last. The MH observation-window was specific to the cohort and defined as the calendar years when the cohort was ascertaining diagnoses for all four MHD. **Study exit** was defined as the earliest date the cohort stopped observing patients, MH observation-window close date, the date the patient was lost to follow-up (defined as 1.5 years with no CD4 or HIV RNA measurement), date of death, or December 31, 2018, whichever came first.

Pearson chi-square tests and Kruskal-Wallis tests were used to assess the differences in demographic and clinical characteristics by each MHD. Annual prevalence for each MHD was estimated and trends were evaluated with a log binomial model with generalized estimating equations (GEE) (as participants could contribute information to multiple years) that included calendar year as a continuous variable to test the hypothesis that there was no change in prevalence from one year to the next.

Retention in care and HIV viral suppression were estimated using data restricted to the most recent years available (Jan 1, 2016, to Dec 31, 2018) and stratified by the four MHD categories and MH multimorbidity. Log binomial models with GEE estimated crude (PR) and adjusted prevalence ratios (aPR) and associated 95% confidence intervals ([-]) for each MHD. Multivariable regression models included age, sex, race/ethnicity, HIV acquisition risk group, and cohort. Subgroup analyses were conducted to evaluate whether having an untreated BD or schizophrenia was associated with a difference on retention in care or viral suppression, compared to no schizophrenia or BD diagnosis. All statistical analyses were conducted using SAS version 9.4 (SAS Institute Inc., Cary North Carolina).

## RESULTS

### Characteristics of the Study Population

Among 122,896 PWH included in this study between 2008-2018, 67,643 (55%) were diagnosed with at least one of the four assessed MHD: 47,553 (39%) were diagnosed with depressive disorder, 34,219 (28%) with anxiety disorder, 11,716 (10%) with BD, and 5,022 (5%) with schizophrenia (**Supplement Figure 1**). In the last two years of observation (2016-2018) (n=64,689), the prevalence of depression was 43%, anxiety was 35%, BD was 10% and schizophrenia was 5% (**Figure 1**).

**Figure 1:**
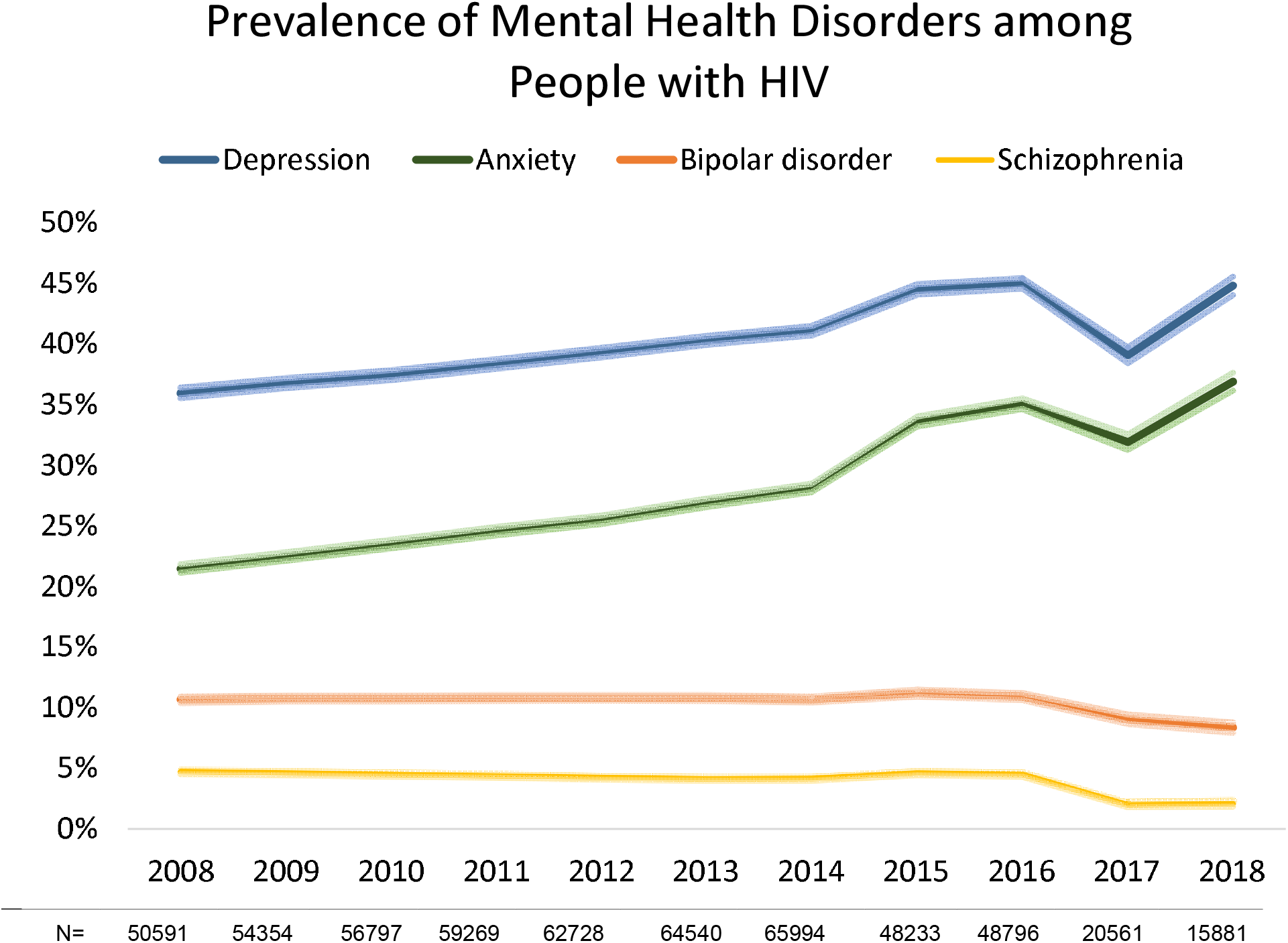
Prevalence (and 95% confidence intervals represented by the lighter color shade) of mental health disorders, 2008-2018. Footnotes: Trends in annual prevalence for each mental health disorder evaluated with a log binomial model with generalized estimating equations, depressive disorder (p for trend= 0.001), anxiety disorder (p for trend= <0.001), bipolar disorder (p for trend= 0.058), and schizophrenia (p for trend= 0.013). Fluctuations in annual prevalence in 2017 and 2018 in mental health disorders may be due to less PWH being observed in our study during this time.

A greater proportion of those with depression, anxiety and BD were non-Hispanic white PWH, whereas a greater proportion of those with schizophrenia were Non-Hispanic Black PWH compared to those without these diagnoses. The median age at study entry was older among those with schizophrenia (51 years [interquartile range (IQR): 44, 57]) compared to those without schizophrenia (46 years [IQR: 37, 54]). PWH with any of these MHD were more likely to have a history of IDU as an HIV acquisition risk factor and were more likely to smoke, have HCV coinfection, and hypertension compared to those without these diagnoses. Those with schizophrenia were more likely to have diabetes, a relationship which was not seen in those with other MHD (**Table 1)**.

**Table 1:** Demographic characteristics at study entry among PWH in the NA-ACCORD and under observation for mental health outcomes between 2008-2018 (N=122,896).

|  | Overall |  | Depression |  |  |  | Anxiety |  |  |  | Bipolar Disorder |  |  |  | Schizophrenia |  |  |  |
| --- | --- | --- | --- | --- | --- | --- | --- | --- | --- | --- | --- | --- | --- | --- | --- | --- | --- | --- |
|  | N=122,896 |  | No Depression<br>n=75,343 |  | Depression<br>n=47,553 |  | No Anxiety<br>n=88,677 |  | Anxiety<br>n=34,219 |  | No Bipolar Disorder<br>n=111,189 |  | Bipolar Disorder<br>n=11,716 |  | No Schizophrenia<br>n=117,874 |  | Schizophrenia<br>n=5,022 |  |
| Characteristics | n | % | n | % | n | % | n | % | n | % | n | % | n | % | N | % | n | % |
| **Age** |  |  |  |  |  |  |  |  |  |  |  |  |  |  |  |  |  |  |
| Median (IQR) | 46 | (37-54) | 45 | (35-53) | 47 | (39-55) | 46 | (36-54) | 47 | (39-54) | 46 | (37-54) | 46 | (38-53) | 46 | (37-54) | 51 | (44-57) |
| **Sex** |  |  |  |  |  |  |  |  |  |  |  |  |  |  |  |  |  |  |
| Male | 104062 | 85% | 63524 | 84% | 40538 | 85% | 74216 | 84% | 29846 | 87% | 94408 | 85% | 9654 | 82% | 99687 | 85% | 4375 | 87% |
| Female | 18832 | 15% | 11819 | 16% | 7013 | 15% | 14458 | 16% | 4374 | 13% | 16770 | 15% | 2062 | 18% | 18184 | 15% | 648 | 13% |
| **Race** |  |  |  |  |  |  |  |  |  |  |  |  |  |  |  |  |  |  |
| Non-Hispanic White | 44816 | 36% | 25796 | 34% | 19019 | 40% | 29022 | 33% | 15794 | 46% | 39626 | 36% | 5190 | 44% | 43516 | 37% | 1300 | 26% |
| Non-Hispanic Black | 52196 | 42% | 32902 | 44% | 19295 | 41% | 40677 | 46% | 11519 | 34% | 47384 | 43% | 4812 | 41% | 49163 | 42% | 3033 | 60% |
| Hispanic | 17651 | 14% | 10938 | 15% | 6713 | 14% | 12557 | 14% | 5094 | 15% | 16382 | 15% | 1269 | 11% | 17100 | 15% | 551 | 11% |
| Non-Hispanic Asian | 2219 | 2% | 1517 | 2% | 703 | 1% | 1714 | 2% | 505 | 1% | 2125 | 2% | 94 | 1% | 2186 | 2% | 33 | 1% |
| Other/Unknown | 6014 | 5% | 4191 | 6% | 1823 | 4% | 4706 | 5% | 1308 | 4% | 5663 | 5% | 351 | 3% | 5908 | 5% | 106 | 2% |
| **HIV acquisition risk** |  |  |  |  |  |  |  |  |  |  |  |  |  |  |  |  |  |  |
| IDU | 21115 | 17% | 10656 | 14% | 10459 | 22% | 12265 | 14% | 8850 | 26% | 16487 | 15% | 4628 | 40% | 18452 | 16% | 2663 | 53% |
| MSM | 42869 | 35% | 28477 | 38% | 14392 | 30% | 32157 | 36% | 10712 | 31% | 39954 | 36% | 2915 | 25% | 42379 | 36% | 490 | 10% |
| Heterosexual contact | 24662 | 20% | 17061 | 23% | 7601 | 16% | 20609 | 23% | 4053 | 12% | 22916 | 21% | 1746 | 15% | 23987 | 20% | 675 | 13% |
| Other/Unknown | 34250 | 28% | 19149 | 25% | 15101 | 32% | 23645 | 27% | 10605 | 31% | 31823 | 29% | 2427 | 21% | 33055 | 28% | 1195 | 24% |
| **Smoking** |  |  |  |  |  |  |  |  |  |  |  |  |  |  |  |  |  |  |
| Never | 26585 | 22% | 15963 | 21% | 10621 | 22% | 18691 | 21% | 7892 | 23% | 24978 | 22% | 1607 | 14% | 25991 | 22% | 594 | 12% |
| Ever | 61191 | 50% | 32229 | 43% | 28963 | 61% | 39501 | 45% | 21690 | 63% | 52673 | 47% | 8518 | 73% | 57253 | 49% | 3938 | 78% |
| Not assessed | 35120 | 29% | 27151 | 36% | 7969 | 17% | 30484 | 34% | 4636 | 14% | 33529 | 30% | 1591 | 14% | 34629 | 29% | 491 | 10% |
| **Hepatitis C infection** | 25009 | 20% | 13459 | 18% | 11551 | 24% | 16323 | 18% | 8686 | 25% | 21006 | 19% | 4003 | 34% | 22814 | 19% | 2195 | 44% |
| **Hepatitis B infection** | 7004 | 6% | 4196 | 6% | 2808* | 6% | 5084 | 6% | 1920* | 6% | 6268 | 6% | 736* | 6% | 6681 | 6% | 323* | 6% |
| **Treated hypertension** | 36543 | 30% | 19099 | 25% | 17445 | 37% | 23625 | 27% | 12918 | 38% | 32221 | 29% | 4322 | 37% | 34030 | 29% | 2513 | 50% |
| **Diabetes** | 12136 | 10% | 6577 | 9% | 5559 | 12% | 8447 | 10% | 3689 | 11% | 10803 | 10% | 1333 | 11% | 11209 | 10% | 927 | 18% |
| CKD stage 2 (eGFR <60) |  |  |  |  |  |  |  |  |  |  |  |  |  |  |  |  |  |  |
| No | 103064 | 84% | 61511 | 82% | 41553 | 87% | 72926 | 82% | 30138 | 88% | 92597 | 83% | 10467 | 89% | 98721 | 84% | 4343 | 86% |
| Yes | 7978 | 6% | 4669 | 6% | 3309 | 7% | 5792 | 7% | 2186 | 6% | 7295 | 7% | 683 | 6% | 7547 | 6% | 431 | 9% |
| Not assessed | 11854 | 10% | 9163 | 12% | 2691 | 6% | 9958 | 11% | 1896 | 6% | 11288 | 10% | 566 | 5% | 11605 | 10% | 249 | 5% |
| Hypercholesterolemia | 26243 | 21% | 14206 | 19% | 12035 | 25% | 17154 | 19% | 9089 | 27% | 23286 | 21% | 2957 | 25% | 24921 | 21% | 1322 | 26% |
| Statin Use | 20674 | 17% | 11003 | 15% | 9671 | 20% | 13625 | 15% | 7049 | 21% | 18631 | 17% | 2043* | 17% | 19608 | 17% | 1066* | 21% |
| CD4 count (cells/mm³) |  |  |  |  |  |  |  |  |  |  |  |  |  |  |  |  |  |  |
| Median (IQR) | 431 (251-634) |  | 428 (247-630) |  | 435 (258-643) |  | 421 (240-626) |  | 454 (279-657) |  | 430 (250-629) |  | 439 (265-642) |  | 423 (234-631) |  | 412 (237-612) |  |
| <200 | 19986 | 16% | 12521 | 17% | 7465 | 16% | 15293 | 17% | 4693 | 14% | 18317 | 16% | 1669 | 14% | 19212 | 16% | 774 | 15% |
| >=200 | 85238 | 70% | 51917 | 69% | 33321 | 70% | 60584 | 69% | 24654 | 72% | 77174 | 70% | 8064 | 69% | 82142 | 70% | 3096 | 61% |
| Missing | 17571 | 14% | 10905 | 14% | 6727 | 14% | 12799 | 14% | 4873 | 14% | 15689 | 14% | 1983 | 17% | 16519 | 14% | 1153 | 23% |
| HIV RNA (copies/mL) |  |  |  |  |  |  |  |  |  |  |  |  |  |  |  |  |  |  |
| <200 | 43075 | 35% | 26436 | 35% | 16638 | 35% | 31138 | 35% | 11937 | 35% | 39150 | 35% | 3925 | 34% | 41604 | 35% | 1471 | 29% |
| >=200 | 51433 | 42% | 32139 | 43% | 19294 | 41% | 37676 | 42% | 13757 | 40% | 46465 | 42% | 4968 | 42% | 49448 | 42% | 1985 | 40% |
| Missing | 28388 | 23% | 16768 | 22% | 11621 | 24% | 19862 | 22% | 8526 | 25% | 25565 | 23% | 2823 | 24% | 26821 | 23% | 1567 | 31% |
| Year of ART initiation |  |  |  |  |  |  |  |  |  |  |  |  |  |  |  |  |  |  |
| 1996-2004 | 33820 | 28% | 17283 | 23% | 16537 | 35% | 22135 | 25% | 11685 | 35% | 29805 | 26% | 4015 | 34% | 31748 | 27% | 2072 | 41% |
| 2005-2009 | 26728 | 22% | 16041 | 21% | 10687 | 22% | 19484 | 22% | 7244 | 21% | 24120 | 22% | 2608 | 22% | 25761 | 22% | 967 | 19% |
| 2010-2018 | 48644 | 40% | 32385 | 43% | 16259 | 34% | 36322 | 41% | 12322 | 36% | 44853 | 40% | 3791 | 32% | 47515 | 40% | 1129 | 22% |
| Not initiate ART | 13690 | 11% | 9622 | 13% | 4068 | 9% | 10726 | 12% | 2964 | 9% | 12391 | 11% | 1299 | 11% | 12835 | 11% | 855 | 17% |
| HIV treatment regimen at ART initiation |  |  |  |  |  |  |  |  |  |  |  |  |  |  |  |  |  |  |
| NNRTI | 14770 | 12% | 10107 | 13% | 4663 | 10% | 10841 | 12% | 3929 | 12% | 13655 | 12% | 1115 | 10% | 14495 | 12% | 275 | 5% |
| PI | 6609 | 5% | 3913 | 5% | 2696 | 6% | 4689 | 5% | 1920 | 6% | 6023 | 5% | 586 | 5% | 6408 | 5% | 201 | 4% |
| Integrase Inhibitor | 42922 | 35% | 25933 | 35% | 16989 | 36% | 30968 | 35% | 11954 | 35% | 39322 | 36% | 3600 | 31% | 41553 | 35% | 1369 | 27% |
| Other/Unknown | 44343 | 36% | 25402 | 34% | 18941 | 40% | 31021 | 35% | 13322 | 39% | 39266 | 35% | 5077 | 43% | 42029 | 36% | 2314 | 46% |
| Never on HAART | 13690 | 11% | 9622 | 13% | 4068 | 9% | 10726 | 12% | 2964 | 9% | 12391 | 11% | 1299 | 11% | 12835 | 11% | 855 | 17% |

P-values calculated using chi-squared tests for all categorical or binary variables and t-tests for all continuous variables. All p-values are <0.001, unless specified with (\*). Due to large numbers of participants statistical significance was common, however clinical significance was considered as a difference in 5% between comparison groups and these differences are bolded.
Age, sex, race/ethnicity, and HIV transmission acquisition group are time-fixed variables, measured at enrollment into the NA-ACCORD.
IDU: injection drug use, MSM: Men who have sex with men, NNRTI: non-nucleoside reverse transcriptase inhibitor, PI: Protease inhibitor.

PWH with any of these four MHD were more likely to have initiated ART prior to 2005 compared to PWH without each MHD, suggesting exposure to earlier ART and a diagnosis of HIV prior to 2005. There were no significant differences in CD4 count or HIV RNA levels at study entry between those with and without depression, anxiety, or BD. PWH with schizophrenia were more likely to have missing CD4 or HIV RNA measurements at study entry and to have never initiated ART (**Table 1**).

### Prevalence of MHD and the effects on the HIV Care Continuum

#### Depression

The prevalence of depressive disorder increased from 35.9% [35.5-36.4%] in 2008 to 44.8% [44.0-45.5%] in 2018 (p=0.001 for annual trend) (**Figure 1**). Those with depression had a higher annual prevalence of retention in care, and lower annual prevalence of viral suppression, compared with those without depression from 2008-2018 (**Figure 2a**). In adjusted models however, PWH with depression had a 2% [0.97-0.98] lower prevalence of retention in care and a statistically nonsignificant 1% [1.00-1.02] higher prevalence of viral suppression than those without depression in recent years (2016-2018) (**Table 2**).

**Table 2:** The impact of Mental Health Disorders on the HIV care continuum among PWH on ART (2016-2018)

|  | n | % | PR<sup>a</sup> | 95% CI | aPR<sup>b</sup> | 95% CI |
| --- | --- | --- | --- | --- | --- | --- |
| Retained in Care (N=54,606) |  |  |  |  |  |  |
| No depression | 30463 | 56% | 1.00 |  | 1.00 |  |
| Depression | 24143 | 44% | **1.03** | **1.03-1.04** | **0.98** | **0.97-0.98** |
| No anxiety | 35068 | 64% | 1.00 |  | 1.00 |  |
| Anxiety | 19538 | 36% | **1.03** | **1.02-1.03** | **0.98** | **0.97-0.98** |
| No bipolar disorder | 48943 | 90% | 1.00 |  | 1.00 |  |
| Bipolar disorder | 5663 | 10% | 1.01 | 1.00-1.01 | 1.00 | 1.00-1.01 |
| No schizophrenia | 52304 | 96% | 1.00 |  | 1.00 |  |
| Schizophrenia | 2302 | 4% | 1.01 | 1.00-1.03 | 1.00 | 1.00- 1.01 |
| No MH Disorder | 20724 | 38% | 1.00 |  | 1.00 |  |
| One MH Disorder | 17101 | 31% | **1.02** | **1.02-1.03** | **1.01** | **1.01- 1.01** |
| MH Multimorbidity<sup>c</sup> | 16781 | 31% | **1.04** | **1.04-1.05** | **1.02** | **1.02- 1.02** |
| Suppressed HIV RNA (≤200 copies/mL) (N=20,231) |  |  |  |  |  |  |
| No depression | 12625 | 62% | 1.00 |  | 1.00 |  |
| Depression | 7606 | 38% | **0.99** | **0.98- 0.99** | 1.01 | 1.00-1.02 |
| No anxiety | 13901 | 69% | 1.00 |  | 1.00 |  |
| Anxiety | 6330 | 31% | 1.00 | 1.00- 1.01 | 1.00 | 1.00-1.01 |
| No bipolar disorder | 18572 | 92% | 1.00 |  | 1.00 |  |
| Bipolar disorder | 1659 | 8% | **0.95** | **0.94-0.97** | **0.98** | **0.98-0.99** |
| No schizophrenia | 19865 | 98% | 1.00 |  | 1.00 |  |
| Schizophrenia | 366 | 2% | **0.96** | **0.93-0.99** | 1.00 | 0.98-1.01 |
| No MH Disorder | 9136 | 45% | 1.00 |  | 1.00 |  |
| One MH Disorder | 6282 | 31% | **0.97** | **0.97-0.98** | 0.99 | 0.99-1.00 |
| MH Multimorbidity<sup>c</sup> | 4813 | 24% | **0.98** | **0.97-0.99** | **0.99** | **0.99-1.00** |

<sup>a</sup> PR = unadjusted prevalence ratio from a log binomial regression model
<sup>b</sup> aPR = adjusted prevalence ratio from a log binomial regression model, adjusted for decade of age, race/ethnicity (due to small numbers, Asian/PI was collapsed into the other/unknown category), sex, HIV acquisition risk group, and cohort.
<sup>c</sup> Mental health multimorbidity included those with ≥2 MH comorbidities
Values with P<0.05 are bolded.

**Figure 2:**
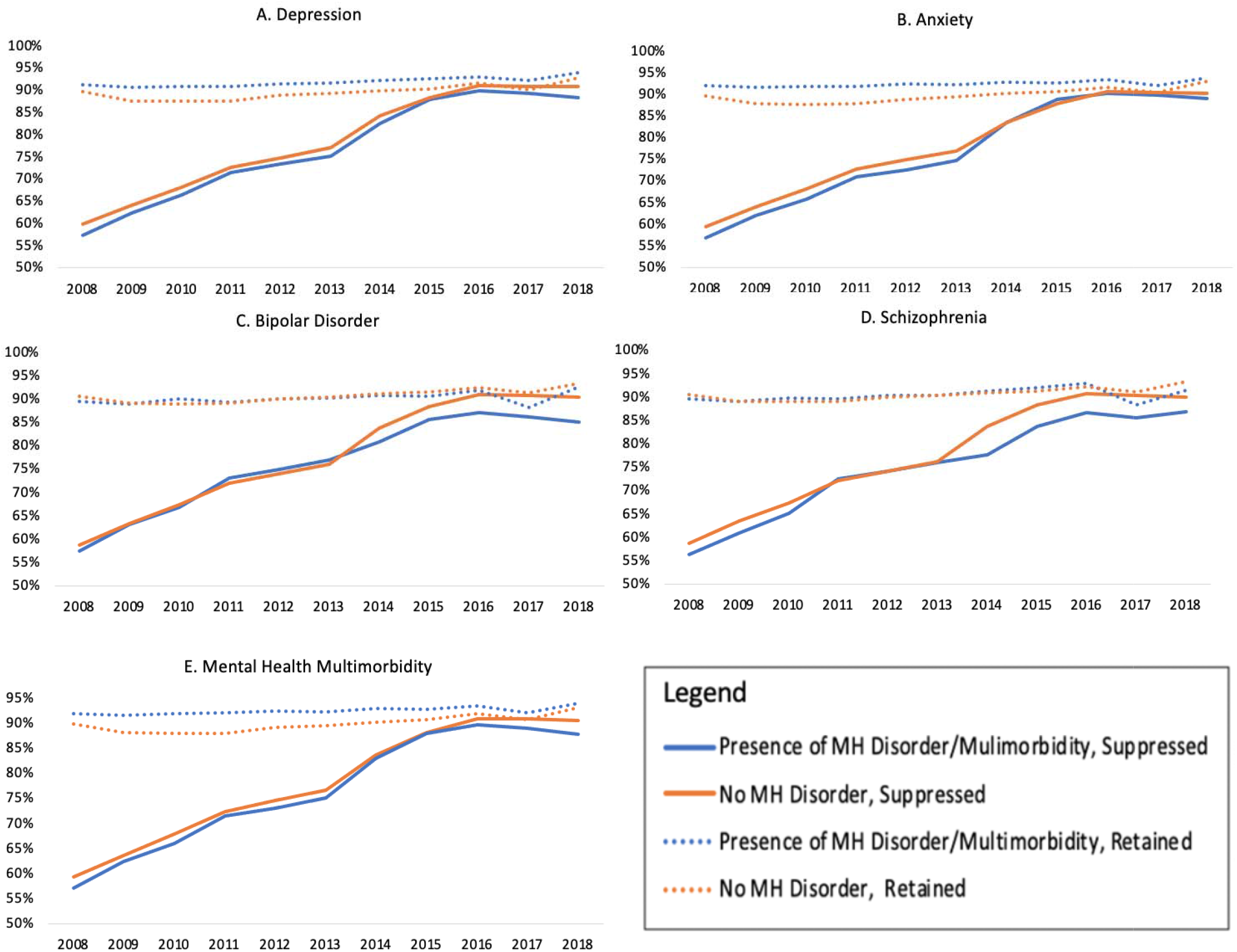
Trends in retention in care and viral suppression by a) depression diagnoses, b) anxiety diagnoses, c) bipolar disorder diagnosis, and c) schizophrenia diagnosis, 2008-2018 (N=122,896) Footnotes: Retention is defined as 2 or more HIV care visits between 2016-2018. Suppression defined as having a viral load ≤200 copies/mL between 2016-2018.

#### Anxiety

The prevalence of anxiety disorder increased from 21.5% [21.1-21.8%] in 2008 to 36.9% [36.1-37.6%] in 2018 (p=0.001 for annual trend) (**Figure 1**). Those with anxiety had slightly higher annual proportion of retention in care compared to those without anxiety. From 2008 to 2013, those with anxiety had a consistently lower viral suppression compared to those without anxiety, however viral suppression was similar by anxiety diagnosis from 2014-2018 (**Figure 2b**). In adjusted models PWH with anxiety had a 2% [0.97-0.98] lower prevalence of retention in care than those without anxiety in recent years (2016-2018). Those with anxiety had no difference in the prevalence of viral suppression compared to those without anxiety in both crude and adjusted models (**Table 2**).

#### Bipolar Disorder

Unlike depression and anxiety, the prevalence of BD decreased slightly over calendar time, from 10.7% [10.4-10.9%] in 2008 to 8.4% [7.9-8.8%] in 2018 (p=0.058 for annual trend) (**Figure 1**). There was no difference in annual trends of the proportions retained in care among those with and without BD from 2008-2018. Those with and without BD had similar proportions virally suppressed from 2008-2013, but from 2014-2018 the proportion suppressed was lower in those with BD compared to those without BD (**Figure 2c**). Those with BD had no difference in the prevalence of retention in care compared to those without (2016-2018) in adjusted models. Those with BD had a 5% [0.94-0.97] lower prevalence of viral suppression than those without BD; however, this changed to a 2% [0.98-0.99] lower prevalence of viral suppression among those with BD after adjustment for demographic characteristics (**Table 2**).

#### Schizophrenia

The prevalence of schizophrenia declined over time, from 4.7% [4.5%-4.9%] in 2008 to 2.1% [1.9%-2.3%] in 2018 (p=0.013 for annual trend) (**Figure 1**). Those with schizophrenia had similar retention in care to those without from 2008-2013, and a higher proportion retained in care from 2014-2018. Those with and without schizophrenia had similar viral suppression until 2013; from 2014-2018 those with schizophrenia were less likely to be virally suppressed compared to those without schizophrenia (**Figure 2d**). In adjusted models PWH with schizophrenia had no difference in the prevalence of retention in care compared to those without schizophrenia (2016-2018). Those with schizophrenia had a 4% [0.93-0.99] decrease in the prevalence of viral suppression compared to those without schizophrenia, and this attenuated to no difference after adjustment for demographic characteristics (**Table 2**).

#### Multimorbidity of MHD among PWH

From 2008-2018, MH multimorbidity prevalence was 24% (29,243/122,896), with 92% of those with schizophrenia (n=5,022), 82% with anxiety (n=34,219), 69% with BD (n=11,716) and 53% with depression (n=47,553) having a diagnosis of at least one other MHD (**Figure 3**). Anxiety was the most common MH comorbidity among those with depression, BD, and schizophrenia. From 2008-2018, the annual proportion of PWH retained in care was similar but consistently higher in those MH multimorbidity compared to those without, whereas viral suppression was similar but consistently lower among PWH with MH multimorbidity, with a gap beginning to widen from 2016-2018 (**Figure 2e**).

**Figure 3:**
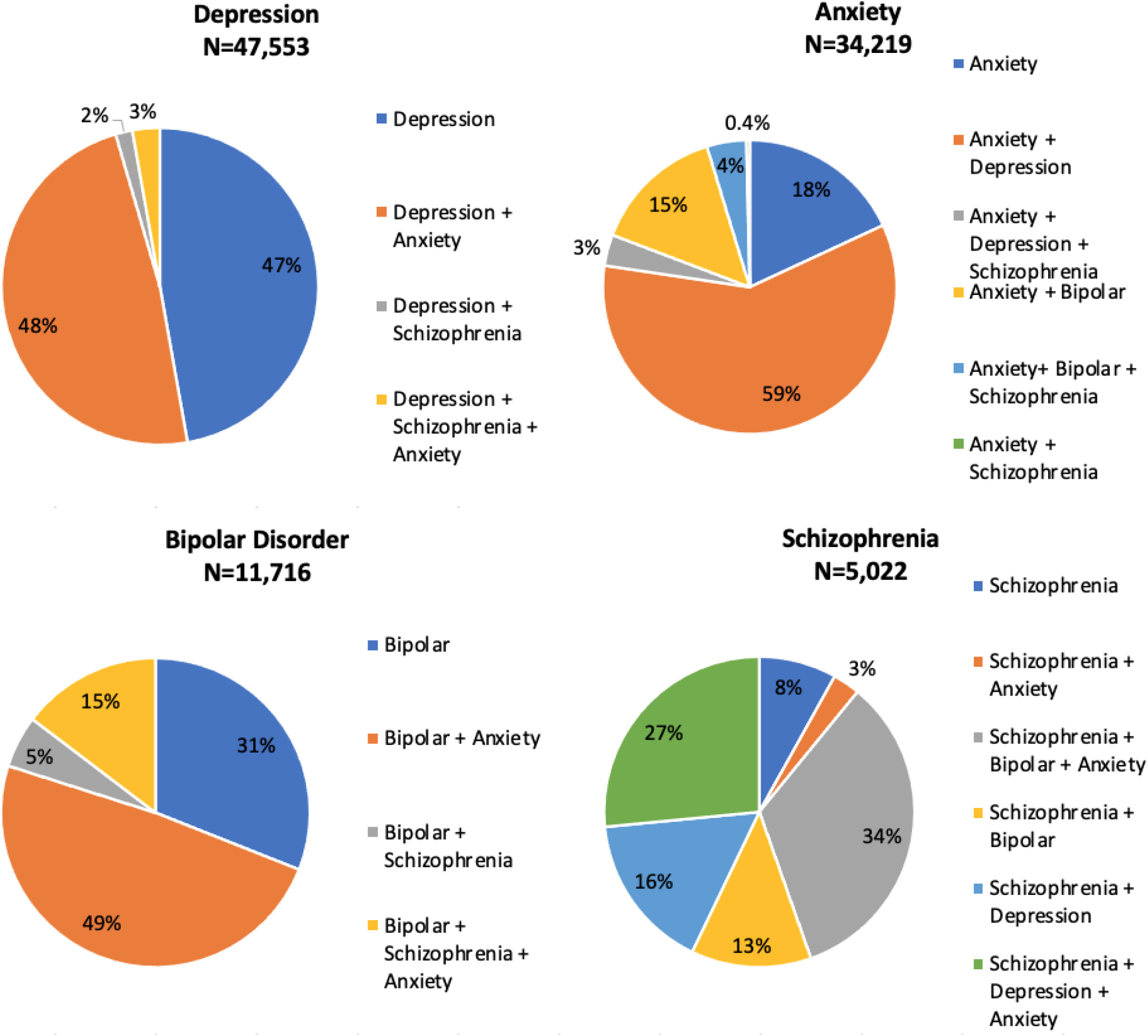
The percentage mental health multimorbidity, by individual mental health comorbidity, 2008-2018. Footnotes: A diagnosis of BD excluded classification of this participant as having depression; these two diagnoses were considered mutually exclusive

In more recent years (2016-2018), the prevalence of MH multimorbidity was 16% (19,457/64,864) among PWH (**Supplement Figure 2**). Those with MH multimorbidity had a 4% [1.04-1.05] higher prevalence of retention in care compared to those without MHD (2016-2018); however, this attenuated to a 2% [1.02-1.02] higher retention in care after adjustment for demographic characteristics. Those with MH multimorbidity had a 2% [0.97-0.99] lower prevalence of viral suppression compared to those without MHD, and this attenuated to a 1% [0.99-1.00] lower prevalence in viral suppression after adjustment for demographic characteristics (**Table 2**).

#### Treated vs. Untreated BD and Schizophrenia

The majority of PWH diagnosed with BD (89%) and schizophrenia (88%) were initiated on mental health treatment. In adjusted analyses there was a statistically nonsignificant 1% lower prevalence of retention in care and viral suppression among PWH with untreated BD compared to those without BD. Untreated schizophrenia was associated with a statistically nonsignificant 1% higher prevalence of retention in care and no difference in viral suppression when compared to PWH without schizophrenia (**Supplemental Table 2**).

## DISCUSSION

The prevalence of MHD is high among PWH in North America, with a large proportion of those with any MHD having MH multimorbidity. MHD disproportionally affect PWH with prevalence rates greater than that reported for the general US population among all four disorders evaluated in this study^[28–30]^. A nationally representative survey of over 9,000 US adults found a lifetime prevalence of 46.4% for any mental health disorder, and a prevalence of 27.7% for two or more disorders^[28]^. We evaluated just four MHD and found 55.1% of PWH had a MHD and 23.8% had ≥ 2 MHD. The general population lifetime prevalence of having a major depressive disorder was 16.6%, generalized anxiety disorder 5.7% and BD was 3.9%^[28]^, compared to PWH in our population with a prevalence of depression being 2.3-fold higher (38.7%), anxiety 4.9-fold higher (27.8%) and bipolar being 2.4-fold higher (9.5%). The median lifetime prevalence of schizophrenia is estimated to be 0.4%^[29]^,^[30, 31]^, and in PWH is 4.1%, over a 10-fold increase.

From 2008-2018, the annual prevalence of depression and anxiety increased among PWH in the NA-ACCORD, with BD and schizophrenia remaining relatively stable until recent years, when prevalence decreased. US General population data suggests that the annual prevalence of depression and anxiety are increasing^[32, 33]^. This increase is likely multifactorial and may be due to a greater recognition over time, increased incidence due to environmental and biologic/genetic factors or increased duration of depression or anxiety disorder due to delayed diagnosis and treatment^[33]^. Temporal trends of the prevalence of BD and schizophrenia in the general population are understudied. However, studies that do report time trends suggest either stable or decreasing prevalence of both BD and schizophrenia^[34–38]^. Reasons for this trend are unknown with suggestions that changing diagnostic criteria may be contributory^[34–37]^. It is possible we are seeing fluctuations in annual prevalence of MHD in 2017 and 2018 due to less PWH being observed in our study during this time.

Although prior studies have demonstrated that PWH with MHD access health services less often and have worse outcomes along the HIV care continuum^[1, 15, 39–41]^, we did not identify large differences associated with MHD or MH multimorbidity on retention in care or viral suppression. However, we did find PWH with depression and anxiety had marginally lower retention in care and those with MH multimorbidity had marginally increased retention in care. Guidance suggests more frequent follow-up visits for PWH with MHD^[42, 43]^, therefore despite small differences observed in retention in care between those with and without MHD, this may signal greater disparity. We found lower viral suppression among PWH with BD and MH multimorbidity compared to those without these diagnoses. Despite these small differences in continuum of care outcomes identified among PWH with MHD and MH multimorbidity, the high prevalence of these conditions among PWH requires ongoing vigilance and tailored interventions to achieve the “Ending the HIV Epidemic” goals of viral suppression^[44]^.

Regardless of MHD diagnosis, the proportion of PWH in NA-ACCORD achieving viral suppression increased between 2008-2018. However, the proportion achieving viral suppression was lower among PWH with BD (vs. no BD) and schizophrenia (vs. no schizophrenia), particularly during the Treat-All era even though retention in care was similar among those with (vs. without) these MHD. This suggests that barriers to viral suppression exist beyond barriers to retention in care which have been exacerbated in the Treat-All era.

PWH with MHD may have higher retention in HIV care and ART adherence if their MH care visits are combined with their HIV care visits, providing convenience and increased access to both types of care^[45–48]^. When evaluating PWH with BD and schizophrenia, we found the majority (~90%) had initiated therapy for these disorders. This may explain the small differences in HIV care continuum among those with MHD vs those without. Our findings highlight the success of HIV care programs to provide PWH with both HIV and MH treatment, but also the ongoing need for improved resources for MH screening, linkage to treatment, and support programs to eliminate the identified gaps. The importance of MH services being integrated into HIV care programs has been emphasized through the COVID-19 pandemic as reports suggest increasing prevalence and worsening symptoms of MHD^[49, 50]^.

One strength of this work is the use of a large, diverse, and representative cohort of PWH who have linked into HIV care in North America. However, enrollment criteria into the NA-ACCORD includes only individuals successfully linked into care. Those who have never linked into care may be more likely to also have MHD, greater severity of MHD or untreated MHD and would not be included in this study population. A second strength lies in the ascertainment for MH diagnoses to classify PWH as ever having a MHD or multimorbidity. However, ascertainment of MHD depended on PWH having been diagnosed with MHD and underdiagnosis is common.

Further limitations include that we did not have sufficient data to evaluate time-varying severity or management of these MHD and are therefore likely identifying a heterogenous population of PWH with MHD. Many of the NA-ACCORD HIV clinical cohort sites provide in-clinic MH screening and treatment; the generalizability of increased retention in care among PWH with MHD and multimorbidity may be limited to clinics with such services. Continuum of care outcome definitions were chosen to be comparable to other research in this field, however it is possible that the sensitivity of these definitions led to the small differences seen between groups. Retention in care was measured with in-person visits and may have been under ascertained if only virtual care visits were attended. Despite this being a large evaluation of MHD in PWH, we were constrained by a small sample size for certain analyses. Finally, there is likely to be residual confounding impacting the associations with HIV care continuum outcomes, particularly by active drug or alcohol use, socioeconomic status, family history of MHD or high stress/traumatic life events, which were not measured.

## Data Availability

Complete data for this study cannot be publicly shared because of legal and ethical restrictions. The NA-ACCORD Principals of Collaboration requires submission and approval of a concept sheet that describes the intended research project for which data are being requested. The NA-ACCORD Executive Committee and the Steering Committee (composed of principle investigators of contributing cohorts) must approve the concept sheet and elect to have their data included for the research project. A signed Data User Agreement is required before data can be released. Guidance for how to obtain NA-ACCORD data are outlined on the NA-ACCORD website (www.naaccord.org/collaboration-policies).

## CONCLUSIONS

This analysis provides novel insight into the prevalence and HIV care continuum outcomes associated with MHD and multimorbidity in PWH. Due to the high prevalence of MHD and multimorbidity among PWH, care providers must remain vigilant with screening for these disorders and providing effective engagement into MH services when needed. Understanding barriers to viral suppression, and effective interventions to overcome such barriers among PWH with MHD and multimorbidity must continue to be a priority to increase the health and well-being of PWH.

---

**Supplemental Figure 1:**
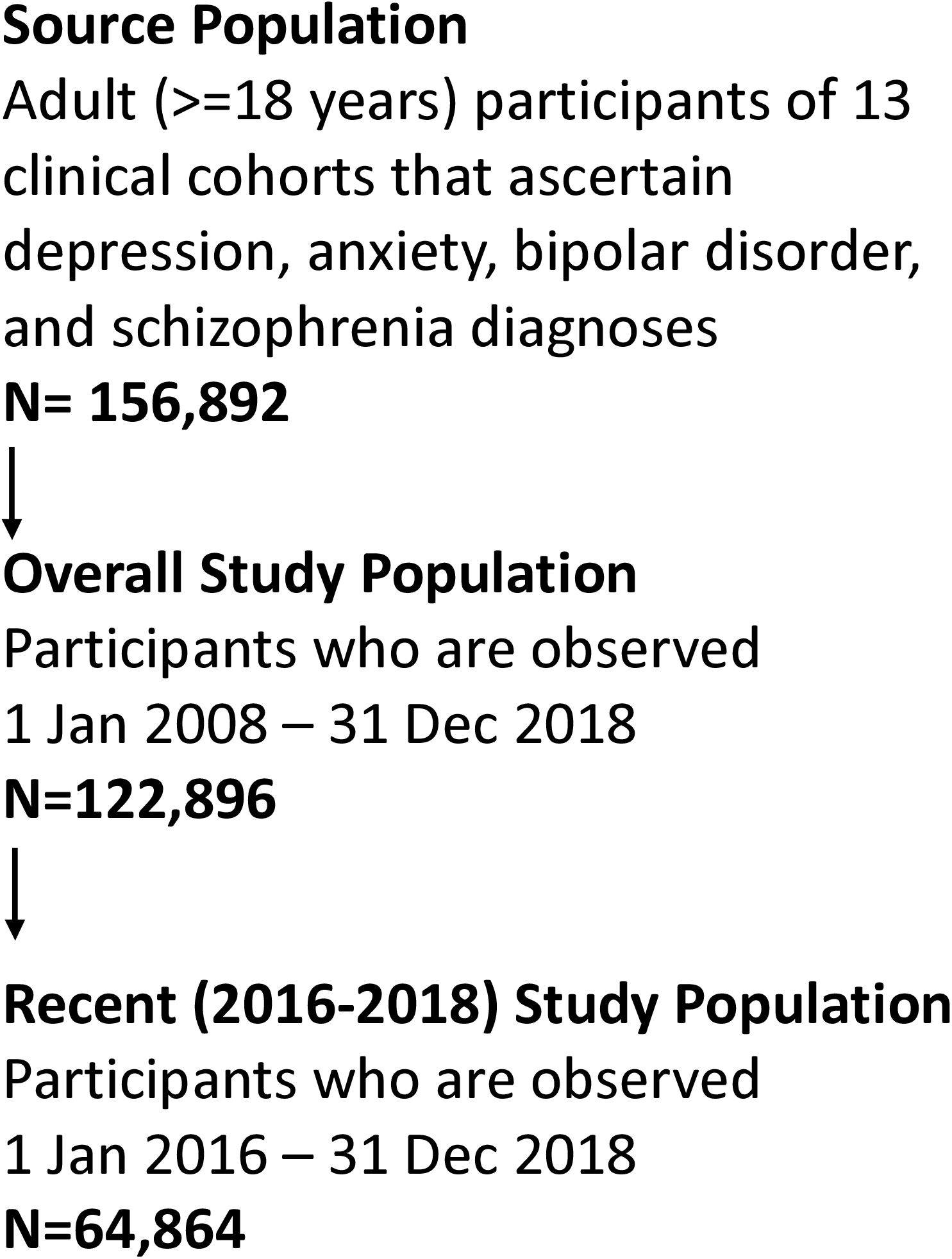
Flow chart of study population selection

**Supplemental Figure 2:**
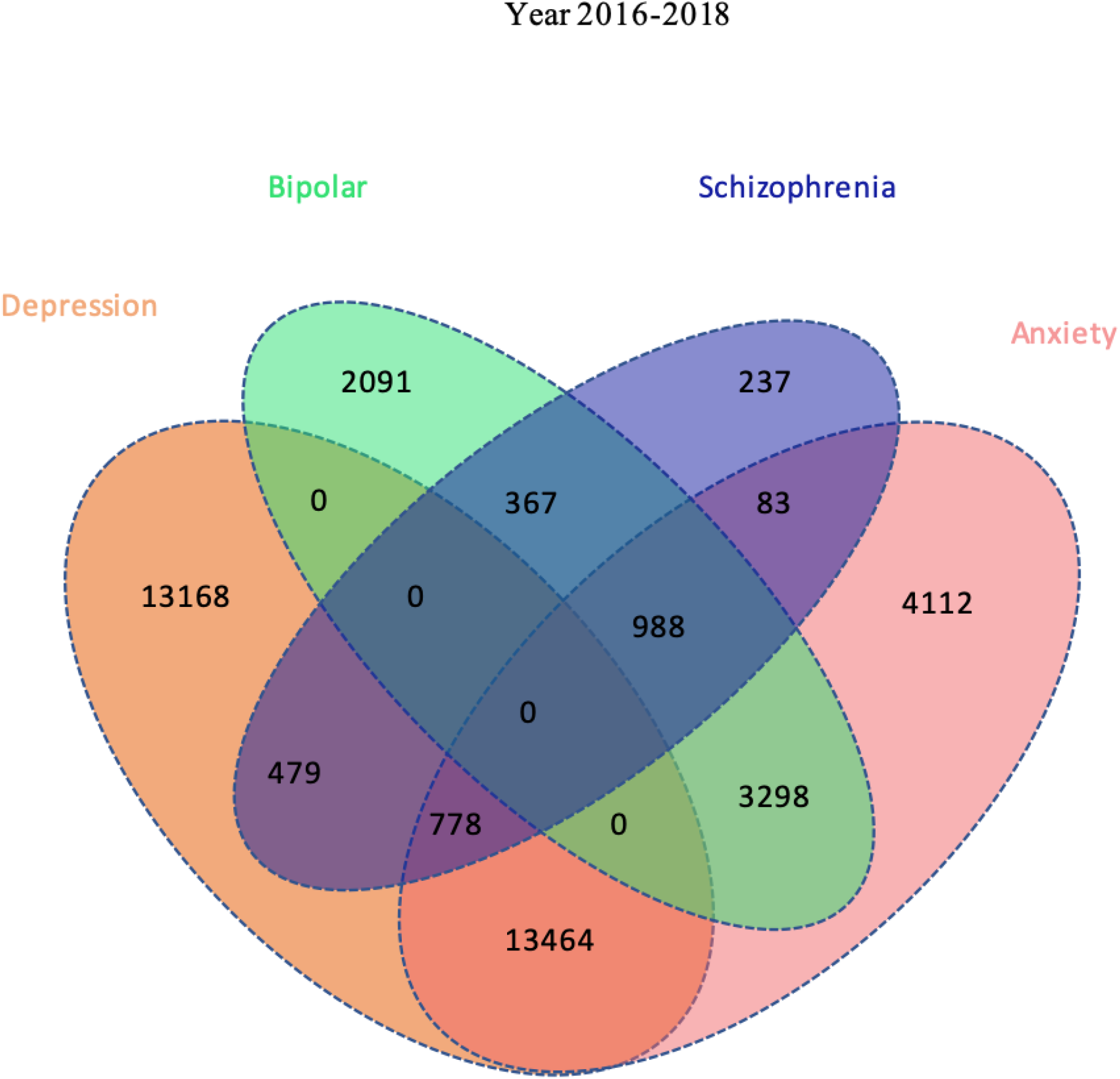
Venn diagram of mental health multimorbidity among people with HIV between 2016-2018 (N=64,864). Footnotes: Depression and bipolar disorder are considered mutually exclusive; therefore, depression diagnoses were excluded among those with bipolar disorder.

**Supplemental Table 1:** Diagnosis codes used to identify Mental health disorders and Medications used to indicate treatment.

| MENTAL HEALTH DISORDER | DIAGNOSIS CODE | DIAGNOSIS CONDITION NAME | MEDICATIONS<sup>A</sup> |
| --- | --- | --- | --- |
| DEPRESSIVE DISORDER | 309.0, F43.21 | ADJUSTMENT DISORDER AND DEPRESSION | NOT ASSESSED |
|  | 296,<br>296.2-296.36,<br>296.82, 296.9,<br>300.4, 309.1,<br>311,<br>F32.0-F32.2,<br>F32.4-F32.5,<br>F32.8,<br>F32.89-F33.2,<br>F33.4-F44.42,<br>F33.8-F34.1,<br>F34.8-F34.9,<br>F39, R45.851,<br>T14.91,<br>T43.592A,<br>T50.902A,<br>V11.1, V62.84,<br>Z91.5 | DEPRESSION |  |
|  | 296.24, 296.34,<br>298.0, F32.3,<br>F33.3 | DEPRESSION AND PSYCHOSIS |  |
| ANXIETY | 309.24, F43.22 | ADJUSTMENT DISORDER AND ANXIETY | NOT ASSESSED |
|  | 293.84-293.127 | ANXIETY DISORDER |  |
| BIPOLAR DISORDER | 296.0-296.16,<br>296.4-296.81,<br>296.89, F30.10-F30.11, F30.3,<br>F30.4, F30.8-F31.13, F31.30-F31.4, F31.60-F31.63, F31.7-F31.9 | BIPOLAR DISORDER | ALPRAZOLAM, ARIPIPRAZOLE, ASENAPINE, BREXIPRAZOLE, CARBAMAZEPINE, CARIPRAZINE, CLOZAPINE, CLONAZEPAM, DIAZEPAM, DIVALPROEX, GABAPENTIN, LAMOTRIGINE, LITHIUM, LORAZEPAM, LURASIDONE, OLANZAPINE, OXCARBAZEPINE, QUETIAPINE, RISPERIDONE, TOPIRAMATE, VALPROIC ACID, ZIPRASIDONE |
|  | 296.04, 296.14, 296.44, 296.54, 296.64, F30.2, F31.12, F31.2, F31.5, F31.64 | BIPOLAR AND PSYCHOSIS |  |
| SCHIZOPHRENIA | 295.0-295.65, 295.08-295.95, F20.0-F20.3, F20.5, F20.81, F20.89, F20.9 | SCHIZOPHRENIA | AMISULPRIDE, AMOXAPINE, ARIPIPRAZOLE, ASENAPINE, BREXPIPIRAZOLE, CARIPRAZINE, CARIPRAZINE HCL, CHLORPROMAZINE, CLOZAPINE, FLUPHENAZINE, HALOPERIDOL, ILOPERIDONE, LOXAPINE, LURASIDONE, MOLINDONE, OLANZAPINE, PALIPERIDONE, PERPHENAZINE, PIMOZIDE, QUETIAPINE, RISPERIDONE, THIORIDAZINE, THIOTHIXENE, TRIFLUOPERAZINE, ZIPRASIDONE, ZUCLOPHNTHIXOL, Anti-psychotic unspecified |

<sup>A</sup>These categories are not mutually exclusive, and many medications are not used exclusively for a single condition.

**Supplemental Table 2:**
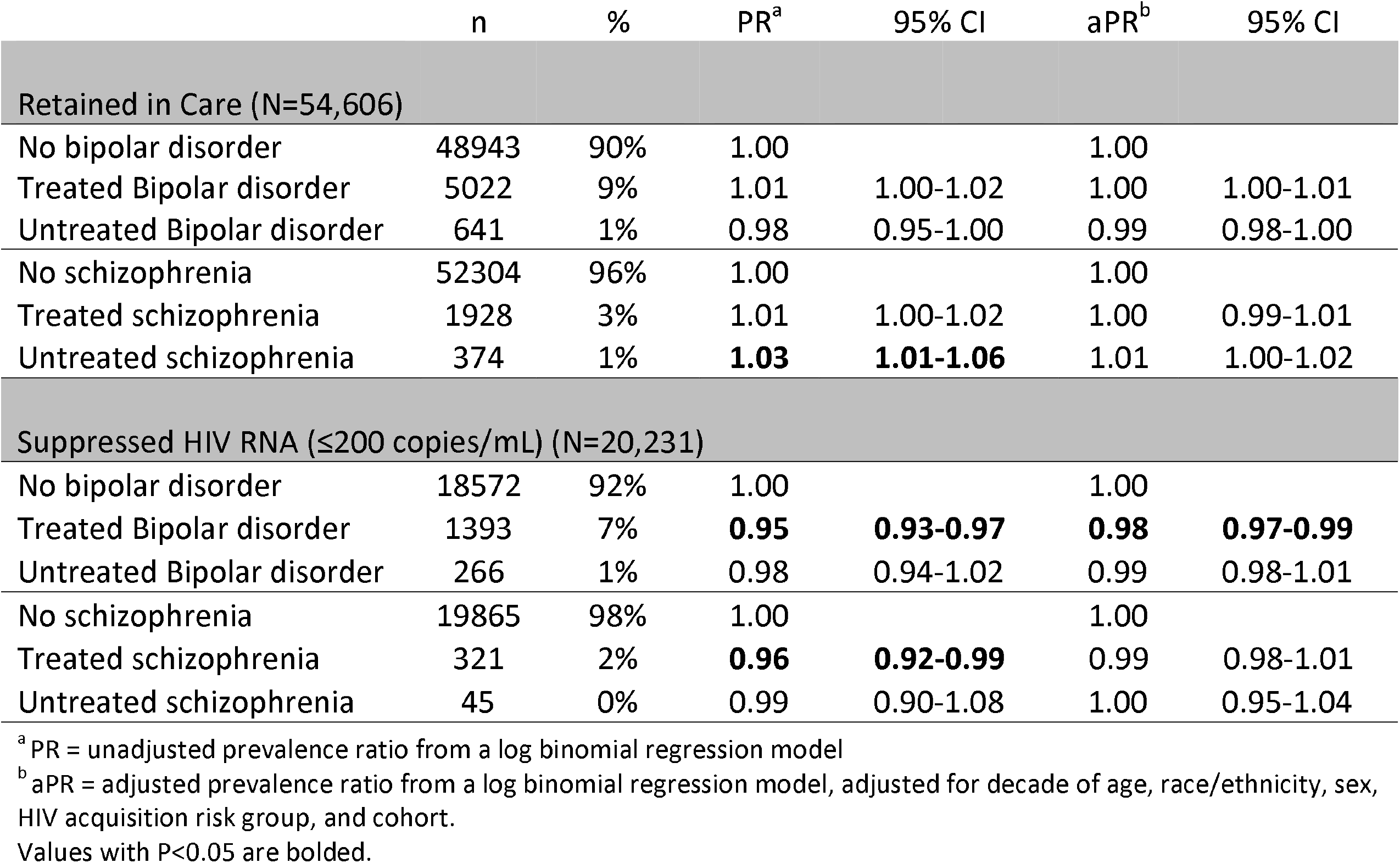
The impact of Mental Health Disorders on the HIV care continuum among PWH on ART (2016-2018)

